# A mixed-methods study of risk factors and experiences of healthcare workers tested for the novel coronavirus in Canada

**DOI:** 10.1101/2021.12.04.21267231

**Authors:** Arnold Ikedichi Okpani, Stephen Barker, Karen Lockhart, Jennifer Grant, Jorge Andrés Delgado-Ron, Muzimkhulu Zungu, Nisha Naicker, Rodney Ehrlich, Annalee Yassi

## Abstract

**Objectives:** We aimed to investigate the contribution of occupational and non-work-related factors to the risk of novel coronavirus (SARS-CoV-2) infection among healthcare workers (HCWs) in Vancouver Coastal Health, British Columbia, Canada. We also aimed to examine how HCWs described their experiences.

**Methods:** We conducted a matched case-control study using data from online and phone questionnaires with optional open-ended questions completed by HCWs who sought SARS-CoV-2 testing between March 2020 and March 2021. Conditional logistic regression and thematic analysis were utilized.

**Results:** Data from 1340 HCWs were included. Free-text responses were provided by 257 respondents. Adjusting for age, gender, race, occupation, and number of weeks since pandemic was declared, community exposure to a known COVID-19 case (adjusted odds ratio -aOR: 2.45; 95% CI 1.67-3.59), and difficulty accessing personal protective equipment -PPE- (aOR: 1.84; 95% CI 1.07-3.17) were associated with higher infection odds. Care-aides/licensed practical nurses had substantially higher risk (aOR: 2.92; 95% CI 1.49-5.70) than medical staff who had the lowest risk. Direct COVID-19 patient care was not associated with elevated risk. HCWs’ experiences reflected the phase of the pandemic when they were tested. Suboptimal communication, mental stress, and situations perceived as unsafe were common sources of dissatisfaction.

**Conclusions:** Community exposures and occupation were important determinants of infection among HCWs in our study. The availability of PPE and clear communication enhanced a sense of safety. Varying levels of risk between occupational groups call for wider targeting of infection prevention measures. Strategies for mitigating community exposure and supporting HCW resilience are required.

## Introduction

The severe acute respiratory syndrome coronavirus 2 (SARS-CoV-2) has had a devastating effect on the health and wellbeing of healthcare workers (HCWs). An estimated 152,888 HCWs had become infected worldwide, with 1,413 dying of the coronavirus disease (COVID-19) by early May 2020.^1^ Infection, disease, and death among HCWs continued through the different phases of the pandemic. HCWs also continue to experience new or worsening mental stress.^2–5^The burden of infection has varied widely across jurisdictions, as has the contribution of known risk factors. In Canada, HCWs accounted for 19.4% of all detected cases between February and July 2020.^6^ By June 2021, that proportion declined to 6.8% of cases, with substantial variation across provinces—12.3% in Quebec, 5.5% in British Columbia (BC), and 4.4% in Ontario.

Some reports indicate that community, rather than the workplace exposure, is the main driver of SARS-CoV-2 infection among HCWs.^7,8^ Others have shown an association between provision of direct care to COVID-19 patients and elevated risk of infection.^9,10^ Further studies found that working in dedicated COVID-19 wards was associated with lower risk, ^11,12^ likely attributable to better availability and use of personal protective equipment (PPE) in units considered high-risk for COVID-19.^13^

The inconsistency of the foregoing findings suggests contextual differences in the predictors of infection risk. The Vancouver Coastal Health (VCH) region of BC, Canada, is an example of a jurisdiction where HCW infections have been comparatively low due to infection prevention and control (IPC) and other mitigation strategies in healthcare.^14^ In addition, VCH implemented other strategies such as asymptomatic onsite testing following suspected or confirmed workplace exposure, dedicated test sites, and priority testing for HCWs. The perception of HCWs of these interventions—in relation to their self-assessed sense of safety at work—has not been documented.

We therefore aimed to investigate the contribution of occupational and non-work-related factors to the risk of SARS-CoV-2 infection among HCWs in VCH, and to explore the lived experience of HCWs who sought testing for SARS-CoV-2, including their perception of personal safety through the phases of the pandemic.

## Methods

VCH is one of BC’s five health authorities and provides care for 25% of the province’s population.^15^ It serves as the referral region for advanced care. Individuals who attended a VCH coronavirus testing centre between March 1, 2020, and March 31, 2021, and self-identified as HCWs, were contacted by the health authority to inform them of a study investigating risk factors for COVID-19 among HCWs. Those who agreed to be contacted were asked to complete a self-administered online questionnaire or to participate in a telephone interview. Each respondent had the opportunity to provide free-text comments on their experience, sense of safety at work and in the community, or any other issues. Data collection took place between November 9, 2020, and June 30, 2021.

We used a mixed-methods approach to integrate qualitative data into a matched-case-control study design. The embedded mixed-method approach^16^ allowed us to examine potential mechanisms that might explain the result of statistical models.

The study protocol was approved by the University of British Columbia Behavioural Research Ethics Board (H20-02517).

### Exposures and variables

We collected data on participants’ demographics, worksite, occupation, activities, and behaviour at and outside of work in the two weeks preceding their test dates. We asked about the respondent’s travel modes to work, exposure to known COVID-19 cases and use of PPE. Respondents tested after December 15, 2020, were asked about vaccination (status and dates). If tested more than once, respondents identified the test for which they had the clearest recall of their activities two weeks prior. With this date and their personal health number—which they supplied as part of the consent process—their test results were extracted from VCH laboratory records.

HCWs who tested positive for SARS-CoV-2 in a nucleic acid amplification test conducted on a nasal swab or mouth gargle specimen were included as cases. HCWs with a negative result were included as controls. Cases and controls were matched without replacement only by the week of the test, in a 1:4 ratio. We excluded individuals with indeterminate or missing test results.

### Quantitative analyses

The odds of a positive SARS-CoV-2 test was estimated for different occupational exposures. These included direct care to COVID-19 patients; exposure to patients’ materials or body fluids; work in proximity (≤2 meters for ≥15 minutes) with colleagues; work with a colleague who subsequently tested positive for SARS-CoV-2 within the two weeks before their own test; and difficulty accessing PPE or reusing PPE. We also estimated the odds of infection for HCWs in non-occupational exposure settings: extended close contact (≤2 meters for ≥15 minutes) with a known COVID-19 infected or symptomatic individual (fever, cough, runny nose, sore throat, shortness of breath); international travel; public transport; and general social interaction with individuals outside of work or home contacts.

We summarized respondents’ characteristics using means, standard deviation, and proportions, stratified by the outcome. Unadjusted logistic regression was used to estimate odds ratios with 95% confidence intervals (CI). Multivariable conditional logistic regression – adjusted for age, gender, race, occupation (where appropriate), and number of weeks since pandemic declared – was used to estimate adjusted odds ratios (aOR). Covariate selection was informed by the results of previous studies.^10,12^

To assess effect modification by pandemic phase, we categorized respondents by test date into three cohorts: respondents tested between March 17, 2020 (date public health emergency declared in BC)^17^, and August 31, 2020 (date BC Provincial Health Officer signaled the start of a potential second wave)^18^, were included in the early cohort (EC). Those tested between September 1 and December 14, 2020, were included in the intermediate cohort (IC). Those who tested between December 15, 2020, when SARS-CoV-2 vaccination was introduced, and March 31, 2021 (study SARS-CoV-2 test eligibility end date), were included in the late cohort (LC).

All statistical analyses were conducted using <u>R version 3.6.1</u>.

### Qualitative analysis

De-identified free-text responses from the questionnaires were exported to a Microsoft Excel spreadsheet for coding and analysis. We first adopted a deductive approach by categorizing responses according to predefined categories. We then examined the data inductively to identify main themes that fit within the preset categories, adding as new categories, main themes that did not fit. Sub-themes were identified within each of the main themes in a constant comparative process. Initial coding was conducted by AO, with 25% independently coded by AD. Disagreements were resolved by consensus.

## Results

We received 1,659 responses to the structured component of the questionnaire, 268 of these being cases. With 1:4 matching, 1,340 observations were included in the quantitative analysis (Figure 1). We received free-text responses to the open-ended component of the questionnaire from 257 respondents. Sixty-one of the responses came from cases and 196 were from controls.

**Figure 1.**
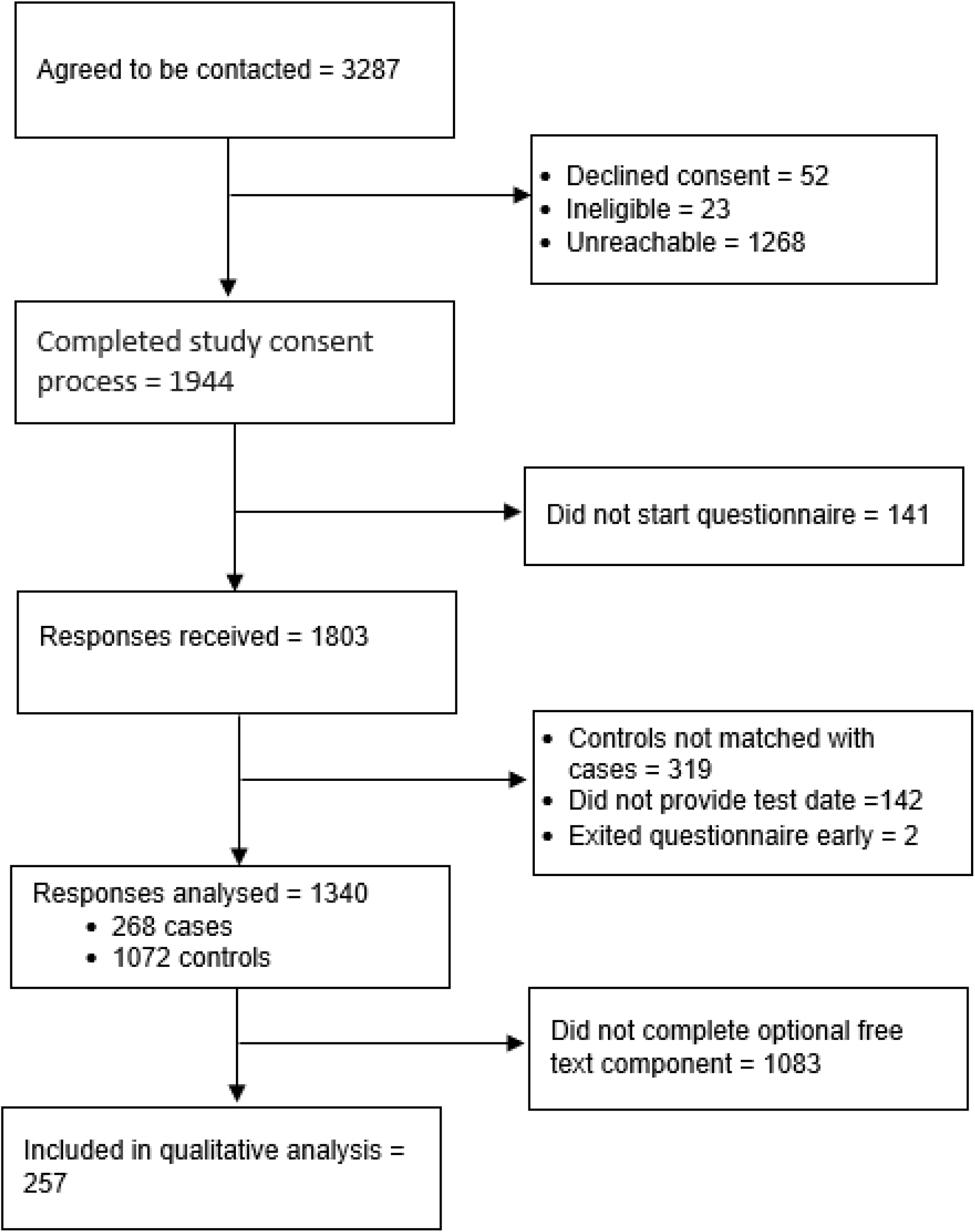
Study flow diagram.

Respondents’ characteristics are shown in Table 1. Most respondents were female, residents of VCH, Canadian citizens, and had tertiary education. Most self-identified as Asian or Non-Hispanic White. There were more acute and community care workers than long-term care (LTC) workers. There was a higher proportion of care-aides/licensed practical nurses (LPNs) among the cases than other occupations.

**Table 1.** Characteristics of study participants. Assessing the relationship between work exposure and Sars-CoV-2 positive test among healthcare workers in Vancouver Coastal Health (March 2020-March 2021)

|  | **Cases (n=268)** | **Controls (n=1072)** |
| --- | --- | --- |
| Mean age, years ± SD | 41.2± 12.9 | 41.3± 11.8 |
| Gender |  |  |
| *Female* | 203 (75.7%) | 816 (76.1%) |
| *Male* | 61 (22.8%) | 241 (25.5%) |
| *Non-binary* | 4 (1.5%) | 10 (0.9%) |
| *Prefer not to answer* | 0 (0.0%) | 5 (0.5%) |
| Postal code of residence |  |  |
| *Vancouver Coastal Health* | 237 (88.4%) | 949 (88.5%) |
| *Fraser Health* | 26 (9.7%) | 114 (10.6%) |
| *Other* | 5 (1.9%) | 9 (0.9%) |
| Status in Canada |  |  |
| *Citizen* | 228 (85.1%) | 972 (90.7%) |
| *Permanent resident* | 30 (11.2%) | 86 (8.0%) |
| *Visitor* | 2 (0.7%) | 5 (0.5%) |
| *Prefer not to answer* | 8 (3.0%) | 9 (0.8%) |
| Indigenous person |  |  |
| *Yes* | 10 (3.7%) | 27 (2.5%) |
| *No* | 257 (95.9%) | 1034 (96.5%) |
| *Prefer not to answer* | 1 (0.4%) | 11 (1.0%) |
| Race |  |  |
| *Asian* | 102 (38.1%) | 344 (32.1%) |
| *Non-Hispanic White* | 128 (47.8%) | 587 (54.8%) |
| *Indigenous* | 7 (2.6%) | 19 (1.8%) |
| *Other* | 23 (8.6%) | 87 (8.1%) |
| *Prefer not to answer* | 8 (3.0%) | 35 (3.3%) |
| Education level |  |  |
| *Secondary or less* | 24 (9.0%) | 78 (7.3%) |
| *Tertiary / University* | 147 (55.1%) | 602 (56.2%) |
| *Postgraduate* | 84 (31.5%) | 355 (33.1%) |
| *Prefer not to answer* | 12 (4.5%) | 37 (3.5%) |
| Work site |  |  |
| *Acute care* | 101 (37.7%) | 470 (43.8%) |
| *Community* | 133 (49.6%) | 473 (44.1%) |
| *Long-term care* | 34 (12.7%) | 129 (12.0%) |
| Underlying health condition |  |  |
| *Yes* | 68 (25.4%) | 257 (24.0%) |
| *No / unknown* | 173 (64.6%) | 710 (66.2%) |
| *No answer* | 27 (10.1%) | 105 (9.8%) |
| Smoking status |  |  |
| *Current smoker* | 5 (1.9%) | 49 (4.6%) |
| *Past smoker* | 47 (17.5%) | 165 (15.4%) |
| *Never smoked* | 189 (70.5%) | 753 (70.2%) |
| *No answer* | 27 (10.1%) | 105 (9.8%) |
| Occupation |  |  |
| *Registered nurses* | 61 (22.8%) | 263 (24.5%) |
| *Care-aides/licensed practical nurses* | 35 (13.1%) | 66 (6.2%) |
| *Administration* | 41 (15.3%) | 152 (14.2%) |
| *Allied health* | 87 (32.5%) | 380 (35.4%) |
| *Medical staff* | 27 (10.1%) | 139 (13.0%) |
| *Support staff* | 9 (3.4%) | 36 (3.4%) |
| *Other/unknown* | 8 (3.0%) | 36 (3.4%) |

SD: standard deviation

Analysis of the unstructured interview indicated that the framing of HCW experiences reflected the phase of the pandemic during which they sought testing. Main themes were lack of clarity and consistency of information – about when to get tested, isolation, contact tracing, test locations and sample collection methods; perceived threats to personal safety at work and in the community; and anxiety about suspected exposure sources.

### Workplace exposures

As shown in Table 2, there was little difference between unadjusted and adjusted odds ratios. In adjusted analysis, provision of direct care to COVID-19 patients was not associated with a positive SARS-CoV-2 test (aOR, 1.05; 95% CI 0.76–1.45). Direct contact with patients’ materials (aOR 1.11; 95% CI 0.82-1.50) and being present for an aerosol-generating procedure (AGP) on a COVID-19 patient (aOR, 1.19; 95% CI 0.59-2.43) were also not associated with higher odds of a positive test. In contrast, HCWs who reported difficulty getting PPE had 1.84 times the odds of infection compared to those who did not report difficulty (95% CI 1.07-3.17). Reuse of PPE was not significantly associated with infection.

**Table 2.** Odds ratio for the relationship between work exposure and SARS-CoV-2 positive test among healthcare workers in Vancouver Coastal Health (March 2020-March 2021)

| Variable | Crude OR (95% CI) | Adjusted OR†<br>(95% CI)<br>n: 268 cases, 1072 controls |
| --- | --- | --- |
| Direct COVID-19 patient care role |  |  |
| *No* | 1 (ref) | 1(ref) |
| *Yes* | 1.04(0.78 - 1.39) | 1.05(0.76 – 1.45) |
| Close contact with known COVID-19 patient |  |  |
| *No / unknown* | 1(ref) | 1(ref) |
| *< 10 times* | 1.11(0.66 - 1.81) | 1.15(0.68 – 1.96) |
| *10 - 50 times* | 1.96(1.07 – 3.45) | 1.67(0.91 – 3.07) |
| *> 50 times* | 2.20(0.88 – 5.15) | 2.12(0.81 – 5.53) |
| Direct contact with patient's materials |  |  |
| *No / unknown* | 1(ref) | 1(ref) |
| *Yes* | 1.17(0.89 - 1.55) | 1.11(0.82 – 1.50) |
| Present for aerosol generating procedure on COVID-19 patient |  |  |
| *No* | 1(ref) | 1(ref) |
| *Yes* | 1.22(0.59 – 2.37) | 1.19(0.59 – 2.43) |
| *Unknown* | 0.88(0.35 – 1.91) | 0.78(0.34 – 1.81) |
| Work site |  |  |
| *Acute care* | 1(ref) | 1(ref) |
| *Community* | 1.31(0.98 – 1.74) | 1.29(0.94 – 1.77) |
| *Long-term care* | 1.22(0.78 – 1.87) | 0.85(0.51 – 1.41) |
| Extended close contact with coworker (within 2m for 15 minutes or more) |  |  |
| *No* | 1(ref) | 1(ref) |
| *Yes* | 0.66(0.50 - 0.87) | 0.65(0.49 – 0.87) |
| Made aware that close-worker contact tested positive afterwards |  |  |
| *No close contact with coworker* | 1(ref) | 1(ref) |
| *Contact, coworker not positive* | 0.72(0.54 – 0.96) | 0.73(0.54 – 0.98) |
| *Contact with positive coworker* | 0.44(0.25 – 0.73) | 0.39(0.23 – 0.68) |
| Work involves contact with patient's materials, belongings, or equipment |  |  |
| *No* | 1(ref) | 1(ref) |
| *Yes* | 0.84(0.60 – 1.17) | 0.80(0.55 – 1.16) |
| *No response* | 0.77(0.49 – 1.19) | 0.70(0.40 – 1.19) |
| Experienced difficulty getting any PPE |  |  |
| *No* | 1(ref) | 1(ref) |
| *Yes* | 1.75(1.05 – 2.83) | 1.84(1.07 – 3.17) |
| Reused PPE on account of an inadequate supply |  |  |
| *No* | 1(ref) | 1(ref) |
| *Yes* | 1.16(0.76 – 1.74) | 1.18(0.77 – 1.80) |

† Adjusted for categorical age, gender, race, occupation, and number of weeks since pandemic declared.
ref=Reference group. PPE: Personal protective equipment.

Several respondents indicated that their employer had worked hard to provide a safe work environment in the face of supply constraints. An IC care-aide thought that:

> *“VCH has really gone above and beyond, getting as much supplies for us as possible. Unfortunately, there is just more demand than what can be supplied. I genuinely feel as though VCH [has] the best intentions for our safety.”*

The adjusted odds ratio of positive SARS-CoV-2 comparing HCWs who worked in close proximity with colleagues with those who didn’t was 0.73 (95% CI 0.54 -0.98) Table 2. The risk of infection was low even if the colleague tested positive in the two weeks before the respondents’ test (aOR, 0.39; 95% CI 0.23-0.68). A LC RN nurse offered a potential explanation for this finding:

> *“[I had been] tested three times, first two times were done due to exposure for a shift, supposedly I possibly worked during period where covid positive coworker worked, so I was asked to be tested twice. I was never symptomatic.”*

As shown in Table 3, comparing infection risk among occupational groups indicated that care-aides/LPNs had 2.73 times the odds of infection compared to medical staff (95% CI 1.47-5.14). Belonging to other occupational groups was not associated with elevated risk. In post-hoc analysis, adjusting for education and home postcode as proxies for socioeconomic status (SES) increased the odds of infection to 2.91, comparing care-aides/LPNs to medical staff.

**Table 3:** Odds ratio for the relationship between occupation and SARS-CoV-2 positive test among healthcare workers in Vancouver Coastal Health region (March 2020-March 2021)

| Variable | Crude OR (95% CI) | Adjusted OR†<br>(95% CI)<br>n: 268 cases, 1072<br>controls | Adjusted OR‡<br>(95% CI)<br>n: 268 cases, 1072<br>controls |
| --- | --- | --- | --- |
| Medical staff | 1(ref) | 1(ref) | 1(ref) |
| Care-aides/licensed practical nurses | 2.73(1.53 – 4.92) | 2.75(1.47 – 5.14) | 2.91(1.47 – 5.77) |
| Administration | 1.39(0.82 – 2.40) | 1.36(0.77 – 2.38) | 1.49(0.81 – 2.73) |
| Allied health | 1.18(0.74 – 1.92) | 1.21(0.73 – 2.00) | 1.28(0.75 – 2.19) |
| Registered nurses | 1.19(0.73 – 1.99) | 1.22(0.72 – 2.09) | 1.37(0.75 – 2.50) |
| Support staff | 1.32(0.55 – 2.98) | 1.17(0.49 – 2.79) | 1.24(0.50 – 3.10) |
| Other/unknown | 1.14(0.45 – 2.64) | 1.00(0.40 – 2.48) | 1.08(0.42 – 2.75) |

ref=Reference group. † Adjusted for categorical age, gender, race, and number of weeks since pandemic declared. ‡ Adjusted for categorical age, gender, race, number of weeks since pandemic declared, education and home postcode. OR: odds ratio

Respondents in work areas that were not considered “high risk” recounted feeling unsafe as they were not prioritized to receive PPE especially when supplies were limited. An EC reception clerk mentioned that:

> *“In March and April there were not enough masks for staff, and everyone thinks clerical workers [do not] touch patients and [they] do not need to wear mask.” An EC cohort administration clerk noted the same concern: “The clinical resource nurse we had refused to make available masks, hand sanitizer, and face shields to clerical staff prior to when I was tested. Even though equipment was available, we had to talk to [them] every time we needed a new mask.”*

### Community exposure

Table 4 shows that exposure to a known COVID-19 case outside of work was associated with infection (aOR, 2.45; 95% CI 1.67-3.59). Similarly, exposure to an individual with symptoms related to COVID-19 was associated with infection (aOR, 1.53; 95% CI 1.07-2.21). International travel, use of public transport, and participation in social interactions were not associated with infection.

**Table 4.**
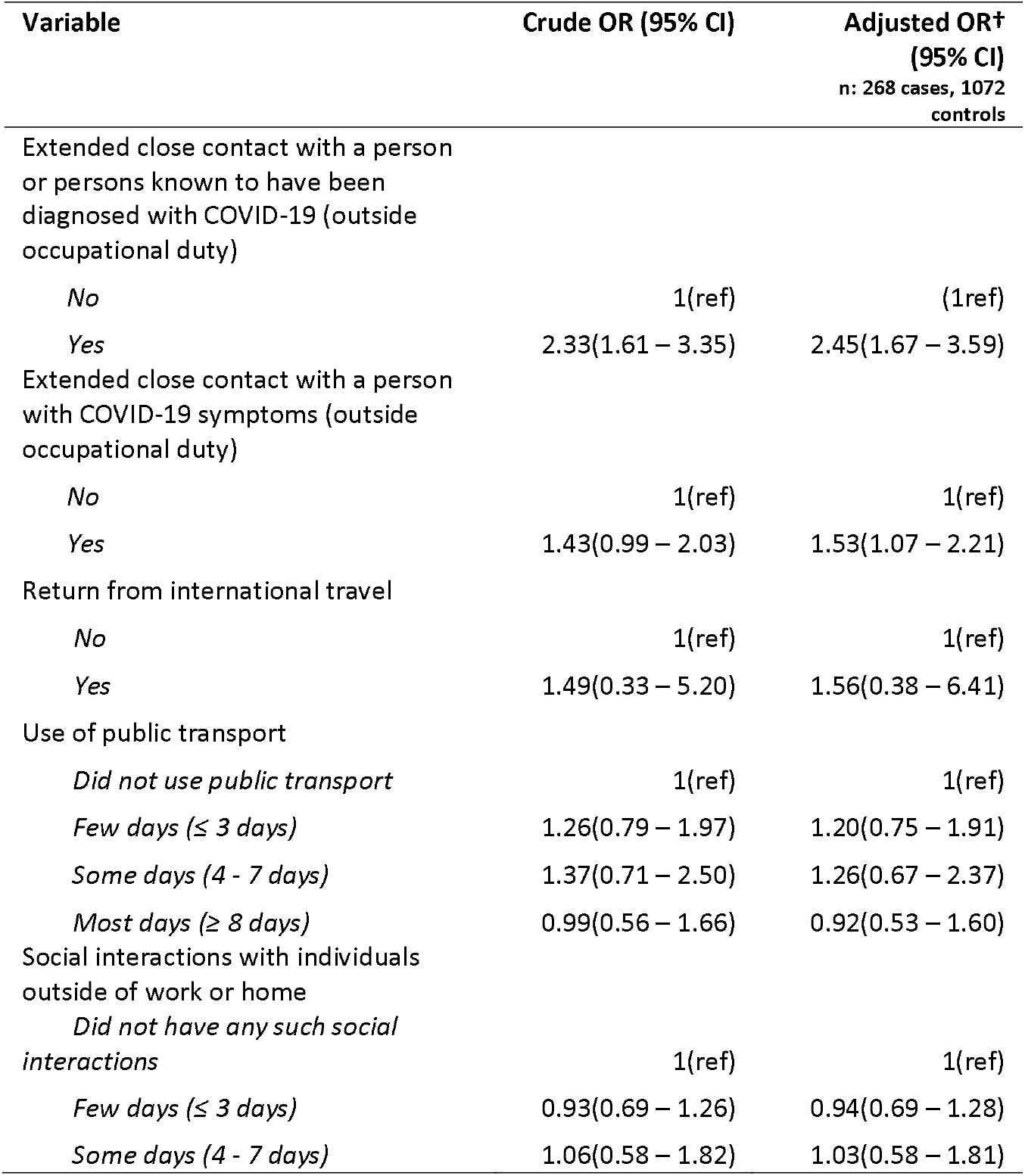
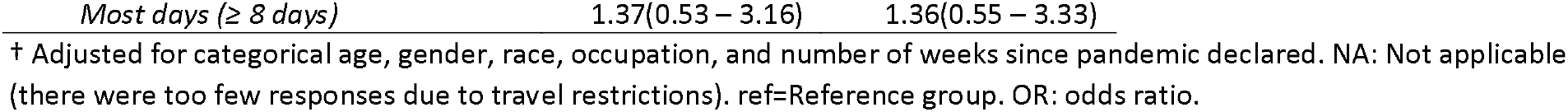
Odds ratio for the relationship between non-work-related risk factors and Sars-CoV-2 positive test among healthcare workers in Vancouver Coastal Health (March 2020-March 2021)

Many HCWs considered themselves safer at work (even when caring for known COVID-19 patients) than in the community. The reason cited was the sense that workplace policies and PPE were keeping them safe whereas they had no control over other people’s choices in the community. A LC RN explained that:

> *“[Our] workplace has taken many precautions to keep us safe. Masks and measures are in place and most people have had at least one dose of vaccine in the office. I do not think it is safe in the community for those unvaccinated or not exposed to virus. “*

A LC social worker had the same view:

> *“[I] Feel safer while at work in hospital, even going on to COVID unit and speaking with patients who are awaiting second negative test, than I do in community (especially on a bus).”*

### Stratified analysis by test date cohorts

As in the main analysis, direct patient care, contact with patients’ materials, being present for an AGP, and worksite were not associated with infection in any of the cohorts. See Supplementary Table 1. The finding of a lower odds of infection among HCWs who worked in close proximity with colleagues was present only in the late test-date cohort, aOR 0.54 (95% CI 0.35-0.82), while the finding of lower odds when a colleague tested positive was present in both the intermediate (aOR, 0.29; 95%CI 0.11-0.77) and late (aOR, 0.30; 95% CI 0.12-0.73) cohorts.

A number of occupational categories showed elevated risk compared to medical staff only in the intermediate cohort: Care-aides/LPNs (aOR, 8.34 (95% CI 2.27-30.61); registered nurses - aOR 4.15 (95% CI 1.27-13.55), and support staff, aOR 6.87 (95% CI 1.27-37.22). See Supplementary Table 2.

International travel, public transit, and social events were not associated with infection, as in the main analysis. Exposure to a known COVID-19 case outside of work was associated with infection only in the intermediate (aOR, 2.73; 95% CI 1.31-5.69), and late (aOR, 2.62; 95% CI 1.51-4.52) test cohorts; See Supplementary Table 3.

### Concerns on getting tested, handling test outcomes, and working during the pandemic

Respondents recounted concerns related to having to deal with rapidly changing policies and protocols, difficulties in accessing tests and results, feelings of stigma, mental stress, and other forms of hardship.

As stated by a LC physical therapist:

> *“My [spouse] and myself kept getting potentially contradictory instructions from multiple different nurses and public health officers, who would also routinely explain that they would need to consult a doctor before they could make further recommendations … our isolation instructions were very poorly communicated. My [spouse] ended up getting 5 COVID tests that were all negative over a one-week period then an antibody test that further confirmed that [they] had not had COVID. An EC registered nurse noted that “the policy at my worksite is unclear and different depending on which manager you speak to. “*

The descriptions of the test experience changed by test site. Onsite testing in health facilities and dedicated lanes for HCWs in community test centres were positively received. HCWs who went to drive-through test sites recounted shorter wait times and fewer privacy concerns than those who went to walk-in sites. An IC unit clerk recounted:

> *“Great having a separate lineup for healthcare workers. Very short time spent waiting in the car via drive-through. Staff were knowledgeable and quick.”*

This contrasted with the experience of an IC registered nurse:

> *“Very little privacy provided at testing site. Personal information was yelled back and forth with other non-health care people around. No attempt was made to make this less open.”*

Some respondents felt they were treated with disdain after they tested positive. Yet others described feeling shame and not wanting to let other know they tested positive. The mental stress of working in the pandemic were also raised. A LC registered nurse recounted their unpleasant experience:

> *“It’s a cold and sad experience and I felt that I was treated as a burden on the system. One person who did the check-in was nasty to me. It was clear that the staff were afraid of me. I certainly never want to go through it again.”*

A LC community support worker shared a similar experience:

> *“I felt extreme stress and shame. All my coworkers found out I had COVID, and some were great about it, but some were not. One thing I feel moving forward is maybe more support for people who are isolating in their rooms. We should also remind people who test positive, it’s not their fault”*.

Recounting the mental stress of isolation, an IC registered nurse stated:

> *“Because of my risk of exposure at work and my underlying condition, I strictly limit my contacts outside of work to my household only and have been doing this the entire pandemic. This is taking a significant toll on my mental health, and on my family as I am unable to be available for support to my elderly and unwell parents.”*

Some respondents indicated they decided to change jobs, worksites, or careers due to anxiety about health and safety. An IC registered nurse recounted their experience before leaving nursing:

> *“Awful situation in LTC facilities. Very limited PPE available. No official announcement on COVID outbreak at facility, heard about it through grapevine. No PPE available at the beginning, letting families in to visit, mixing COVID patients with other patients for dining. Completely run down and very stressed. Subsequently left the role and nursing and now working in another sector.”*

An IC social worker described a similar experience:

> *“I worked at a clinic [where] I felt very unsafe there. The IPAC practices were not good. They let patients in without masks. They had no isolation room. [Three] Workers got Covid at that site, including me. I have resigned from that site.”*

## Discussion

In this study of HCWs in the Vancouver Coastal Health region, we found no association between direct care to COVID-19 patients and a positive SARS-CoV-2 test. However, lack of access to PPE, and working as a care-aide/LPN were predictive of HCW infection. Exposure to a known or suspected case of COVID-19 in the community was a strong predictor of infection.

A previous study by our research group found that rates of infection among VCH HCWs were similar to those of the background population,^14^ reinforcing the importance of measures like PPE, point-of-care risk assessment, contact tracing, and infection prevention and control in keeping HCWs safe. A study by Baker and colleagues,^7^ and another by Jacob and colleagues, ^8^ also found that community exposures were more strongly associated with SARS-CoV-2 seropositivity than workplace exposure. Working in LTC as opposed to acute care has been identified as being associated with infection in other studies (including our previous study). We did not find a significantly higher risk in LTC after accounting for occupation. However, we found that care-aides/LPNs who work mostly in LTC had a higher risk of infection compared to medical staff. This difference in risk is not entirely attributable to SES as we found a stronger association when we accounted for education and home postcode as proxies for SES in the analysis. Residual confounding by SES remains a possibility, however.

Our finding of a much lower risk of infection among HCWs who reported working in proximity with infected colleagues could result from the higher test frequency following exposure, leading to overrepresentation of such contacts among the controls. Public Health protocols in BC promoted comprehensive contact-tracing and a very low testing threshold—including asymptomatic testing—for HCWs exposed at work. The finding could also be partly attributable to better attention to, and availability of, PPE among HCWs in patient-facing settings, as suggested by the lack of association between direct COVID-19 patient care and risk of infection. The foregoing agree with the findings of a rapid review of workplace policies useful in preventing COVID-19.^19^ Furthermore, HCWs in high COVID-19 transmission settings were prioritized for vaccination in BC at a time when vaccines were not widely available.^20^

Notable findings in the period stratified analysis were very high odds ratios relative to medical staff for care-aides/LPNs, registered nurses, and support staff only in the intermediate cohort. Overall, however, findings were imprecise due to low case numbers in each subgroup, limiting any conclusion about trends.

The experiences of mental stress and stigma reported by our study participants are consistent with findings from previous studies.^22–26^ As much as 75% of Canadian HCWs involved in COVID-19 direct care reported that their mental health deteriorated since the start of the pandemic.^5^ Not only has stigmatization of HCWs by certain members of the community been a problem through the pandemic, ^27^ but so has stigmatization by fellow HCWs of colleagues who tested positive.^28^

These reports of mental stress call for specifically targeted strategies to protect HCWs from more than just biological hazards during the COVID-19 pandemic. As the pandemic drags on and new waves of infection continue to strain health services, measures to promote the resilience of weary HCWs are urgently needed. As HCWs have homes to return to each day, research into strategies that can promote HCW safety beyond the workplace is needed.

Acquiring SARS-CoV-2 infection in the workplace is not inevitable. What is required is a reappraisal of the approach to implementing mitigation strategies to ensure that all HCWs – not just those in perceived “high risk” work environments – are equally protected. The higher rate of infection among care-aides/LPNs and HCWs who had difficulty getting PPE calls for better attention to ensuring a universal precautions approach throughout the healthcare sector. This will mean ensuring that the same level of IPC measures, PPE training and supply is available when needed to HCWs regardless of their occupational stature or setting in which they work.

### Limitations

Respondents self-selected into the study. Consequently, participants could be systematically different from HCWs who chose not to participate. Our findings, however, are consistent with the result of our group’s previous study^14^ among VCH HCWs who comprised more than 80% of the respondents in this study. Secondly, in case-control studies, there is potential of differential recall between cases and controls. The method of generation of qualitative data we adopted is a third limitation as the use of optional free-text questions precluded full in-depth interviews. That, however, was not a major objective of this study, and more rigorous exploration of the themes generated would require a separate study.

## Data Availability

All data produced in the present study are available upon reasonable request to the authors

## Acknowledgements

The study protocol was adapted from the WHO *Assessment of risk factors for coronavirus disease 2019 (COVID-19) in health workers: protocol for a case-control study* (reference number WHO/2019-nCoV/HCW_RF_CaseControlProtocol/2020.1) available from https://apps.who.int/iris/handle/10665/332187

We are grateful to the medical student volunteer team at the Physician Occupational Safety and Health (POSH) service of VCH who conducted the recruitment calls. We thank Patricia Gray, Chloe Lim, Olivia Tsai, and Vanessa Wong for conducting the telephone interviews. Special thanks to Amee Manges and Jerry Spiegel of the School of Population and Public Health of UBC for their helpful inputs to the design of this study. We are grateful to members of our research collaboration who provided feedback throughout the stages of design and implementation of this project.

---

**Supplementary Table 1:**
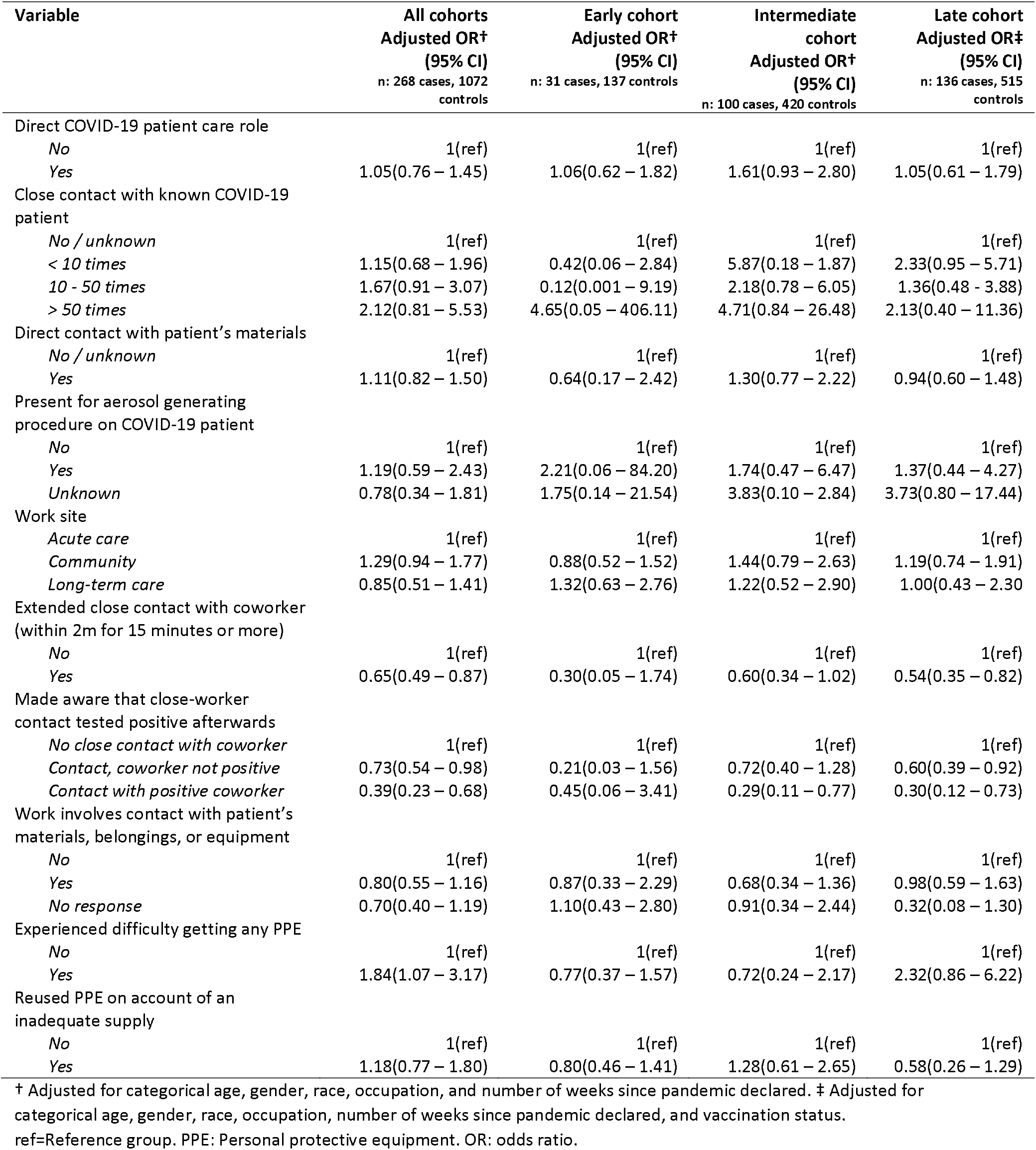
Odds ratios for workplace exposures stratified by test date cohort.

**Supplementary Table 2.** Odds ratios by occupation stratified by test date cohort.

| Variable | All cohorts<br>Adjusted OR†<br>(95% CI)<br>n: 268 cases, 1072 controls | Early cohort<br>Adjusted OR†<br>(95% CI)<br>n: 31 cases, 137 controls | Intermediate cohort<br>Adjusted OR†<br>(95% CI)<br>n: 100 cases, 420 controls | Late cohort<br>Adjusted OR‡<br>(95% CI)<br>n: 136 cases, 515 controls |
| --- | --- | --- | --- | --- |
| Medical staff | 1(ref) | 1(ref) | 1(ref) | 1(ref) |
| Care-aides/licensed practical nurses | 2.75(1.47 – 5.14) | 0.76(0.30 – 1.90) | 8.34(2.27 – 30.61) | 1.86(0.69 – 5.05) |
| Administration | 1.36(0.77 – 2.38) | 1.08(0.44 – 2.65) | 2.44 (0.69 – 8.67) | 1.11(0.48 – 2.56) |
| Allied health | 1.21(0.73 – 2.00) | 0.77(0.35 – 1.66) | 2.27(0.72 – 7.14) | 0.88(0.42–1.82) |
| Registered nurses | 1.22(0.72 – 2.09) | 0.95(0.38 – 2.38) | 4.15(1.27–13.55) | 0.66(0.30 – 1.52) |
| Support staff | 1.17(0.49 – 2.79) | 0.89(0.29 – 2.72) | 6.87(1.27 – 37.22) | 0.67(0.16 – 2.81) |
| Other/unknown | 1.00(0.40 – 2.48) | 2.94(0.33 – 26.33) | 2.10(0.32 – 10.63) | 0.90(0.27 – 3.05) |

ref=Reference group. † Adjusted for categorical age, gender, race, and number of weeks since pandemic declared. ‡ Adjusted for categorical age, gender, race, number of weeks since pandemic declared, and vaccination status. OR: odds ratio

**Supplementary Table 3.** Odds ratio for the relationship between non-work-related risk factors and SARS-CoV-2 positive stratified by test date cohorts.

| Variable | All cohorts<br>Adjusted OR†<br>n: 268 cases, 1072 controls | Early cohort<br>Adjusted OR†<br>n: 31 cases, 137 controls | Intermediate cohort<br>Adjusted OR†<br>n: 100 cases, 420 controls | Late cohort<br>Adjusted OR‡<br>n: 136 cases, 515 controls |
| --- | --- | --- | --- | --- |
| Extended close contact with a person or persons known to have been diagnosed with COVID-19 (outside occupational duty) |  |  |  |  |
| No | (1ref) | (1ref) | 1(ref) | (ref) |
| Yes | 2.45(1.67 – 3.59) | 1.03(0.47 – 2.23) | 2.73(1.31 – 5.69) | 2.62(1.51 – 4.52) |
| Extended close contact with a person with COVID-19 symptoms (outside occupational duty) |  |  |  |  |
| No | 1(ref) | (1ref) | 1(ref) | 1(ref) |
| Yes | 1.53(1.07 – 2.21) | 1.16(0.58 – 2.30) | 1.41(0.76 – 2.64) | 1.63(0.94 – 2.80) |
| Return from international travel |  |  |  |  |
| No | 1(ref) | (1ref) | 1(ref) | 1(ref) |
| Yes | 1.56(0.38 – 6.41) | 1.74(0.51 – 5.87) | NA | 1.84(0.12 – 28.22) |
| Use of public transport |  |  |  |  |
| Did not use public transport | 1(ref) | (1ref) | 1(ref) | 1(ref) |
| Few days ($\leq 3$ days) | 1.20(0.75 – 1.91) | 0.76(0.35 – 1.62) | 1.24(0.53 – 2.91) | 1.08(0.53 – 2.19) |
| Some days (4 - 7 days) | 1.26(0.67 – 2.37) | 0.58(0.19 – 1.78) | 0.66(0.12 – 3.55) | 1.96(0.82 – 4.67) |
| Most days ($\geq 8$ days) | 0.92(0.53 – 1.60) | 1.13(0.60 – 2.13) | 0.60(0.19 – 1.88) | 1.31(0.60 – 2.87) |
| Social interactions with individuals outside of work or home |  |  |  |  |
| Did not have any such social interactions | 1(ref) | (1ref) | 1(ref) | 1(ref) |
| Few days ($\leq 3$ days) | 0.94(0.69 – 1.28) | 1.20(0.73 – 2.00) | 1.48(0.85 – 2.56) | 0.73(0.50 – 1.14) |
| Some days (4 - 7 days) | 1.03(0.58 – 1.81) | 1.02(0.42 – 2.45) | 3.23(1.16 – 9.01) | 0.49(0.19 – 1.26) |
| *Most days ($\geq 8$ days)* | 1.36(0.55 – 3.33) | 0.82(0.19 – 3.53) | 2.55(0.36 – 18.80) | 0.91(0.23 – 3.57) |

† Adjusted for categorical age, gender, race, and number of weeks since pandemic declared. ‡ Adjusted for categorical age, gender, race, number of weeks since pandemic declared, and vaccination status. ¥
NA: Not applicable (there were too few responses due to travel restrictions). ref=Reference group. OR: odds ratio

